# Sweetspot mapping in deep brain stimulation: Strengths and limitations of current approaches

**DOI:** 10.1101/2020.09.08.20190223

**Authors:** Till A. Dembek, Juan Carlos Baldermann, Jan-Niklas Petry-Schmelzer, Hannah Jergas, Harald Treuer, Veerle Visser-Vandewalle, Haidar S. Dafsari, Michael T. Barbe

**Affiliations:** Department of Neurology, Faculty of Medicine, University of Cologne, Cologne, Germany; Department of Stereotactic and Functional Neurosurgery, Faculty of Medicine, University of Cologne, Cologne, Germany

**Keywords:** Deep Brain Stimulation, Probabilistic Mapping, Sweetspot, Voxel-wise statistics

## Abstract

**Objective:** Open questions remain regarding the optimal target, or sweetspot, for deep brain stimulation (DBS) in e.g. Parkinson’s Disease. Previous studies introduced different methods of mapping DBS effects to determine sweetspots. While having a direct impact on surgical targeting and postoperative programming in DBS, these methods so far have not been investigated in ground-truth data.

**Materials & Methods:** This study investigated five previously published DBS mapping methods regarding their potential to correctly identify a ground-truth sweetspot. Methods were investigated *in silico* in eight different use-case scenarios, which incorporated different types of clinical data, noise, and differences in underlying neuroanatomy. Dice-coefficients were calculated to determine the overlap between identified sweetspots and the ground-truth. Additionally, out-of-sample predictive capabilities were assessed using the amount of explained variance R^2^.

**Results:** The five investigated methods resulted in highly variable sweetspots. Methods based on voxel-wise statistics against average outcomes showed the best performance overall. While predictive capabilities were high, even in the best of cases Dice-coefficients remained limited to values around 0.5, highlighting the overall limitations of sweetspot identification.

**Conclusions:** This study highlights the strengths and limitations of current approaches to DBS sweetspot mapping. Those limitations need to be taken into account when considering the clinical implications. All future approaches should be investigated *in silico* before being applied to clinical data.

## Introduction

Deep brain stimulation (DBS) is an established treatment option for a variety of neurological disorders. For example, electric stimulation of the subthalamic nucleus (STN) and its surroundings via implanted electrodes can lead to a marked and lasting improvement of motor^1^ and non-motor symptoms^2^ in Parkinson’s disease (PD). However, DBS still is an evolving therapy with new indications, new surgical targets, and new stimulation strategies and techniques emerging every year^3–5^. To achieve the greatest benefit in each individual patient, electrodes need to be implanted accurately in the optimal position and stimulation parameters have to be finely adjusted to stimulate the exact, most favorable region. Unfortunately, many open questions still remain regarding where these optimal stimulation locations/regions are situated and whether they are comprised of particular neuroanatomical regions i.e. nuclei or fiber tracts^6–8^. Determining optimal stimulation regions thus is a crucial step to move the field of DBS forward and a variety of research groups, employing heterogeneous strategies have made efforts in finding these ‘sweetspots’ for different indications for almost a decade^6,8–16^. In most cases, sweetspots were determined by incorporating different data, consisting of 1) the locations of implanted electrodes as determined from pre- and postoperative neuroimaging, 2) stimulation parameters and models of electrical stimulation, and 3) clinical outcome parameters. By then mapping clinical outcomes to the locations of stimulation over groups of patients and employing different statistical methods or thresholding a sweetspot linked to most advantageous outcome can be determined. Crucially, a variety of such mapping strategies have been proposed but since the ‘real’ sweetspots are unknown, it is difficult to investigate and compare their validity. Additionally, these methods sweetspots have neither been validated in ground truth data nor in prospective clinical trials where these sweetspots are explicitly targeted. While the results of DBS sweetspot-analyses have reached a wide audience and may directly impact current concepts regarding the neuroanatomical origins of DBS effects, surgical DBS targeting, and postoperative DBS programming, it remains unclear whether the methods on which these sweetspots were based are able to produce reliable results. The aim of this study is to address these dilemmas by comparing different mapping strategies *in silico*, using a ‘ground-truth’ sweetspot. In doing so, we aim to give a comprehensive overview about the strengths and limitations in existing strategies for sweetspot-mapping in DBS as well as to provide guidelines to achieve more accurate and comparable mapping results for future DBS trials.

## Methods

### Ethics & informed consent

This was an *in silico* trial which used no clinical data except retrospectively gathered, fully anonymized locations of DBS leads. Consecutively no ethics vote or informed consent was needed for this study.

### Dataset

The dataset was based on electrode locations from n = 100 PD patients who had been bilaterally implanted with DBS electrodes targeting the STN in our center. Electrode locations had been determined using the Lead-DBS toolbox. Preoperative MRI and postoperative CT images were coregistered linearly using Advanced Normalization Tools (ANTs).^19^ Images were then normalized into the MNI space (2009b, nonlinear, asymmetric) using the symmetric diffeomorphic registration approach (SyN) implemented in ANTs and the ‘subcortical refine’ setting as implemented in Lead-DBS.^19–21^ Leads were identified using the PaCER algorithm.^22^ The Medtronic 3389 lead model was chosen for all patients. For further analysis, all leads were transferred to the right hemisphere. All steps were visually inspected by an experienced user (TAD).

### Stimulation volumes

For each lead we calculated binarized stimulation volumes (or volumes of tissue activated, VTA). VTAs were estimated using the FastField algorithm as implemented in Lead-DBS using an electric-field threshold of 0.2 V/mm and an isotropic conductivity of 0.1 S/m^23,24^. VTAs were calculated for each electrode on each lead with amplitudes ranging from 1 to 5 mA in 1 mA steps and a pulsewidth of 60 μs. This resulted in 5 VTAs per electrode, 20 VTAs per lead and 4000 VTAs over all 100 patients. All VTAs were transferred to a 100×100×100 isotropic voxel-space with a voxel-size of 0.2×0.2×0.2 mm which covered the whole STN area.

### ‘Ground-truth’ sweetspot and clinical improvement

For this *in silico* study the sensorimotor subpart of the STN, as defined in the DISTAL atlas,^25^ was defined as the ‘ground-truth’ sweetspot. We presumed that a higher stimulation overlap with the sensorimotor STN would be the sole predictor of ‘clinical’ outcome. A VTA which showed no overlap with the sensorimotor STN would thus lead to an improvement of 0% while a VTA covering the whole sensorimotor STN would lead to an improvement of 100%. In reality however, clinical outcome is of course influenced by a variety of other factors and often only depicted on low-resolution clinical scales. To address this issue, we also included an analysis where we added Gaussian noise to the calculated improvements based on their variance. Additionally, we conducted a separate analysis where we added the neighboring internal capsule (1C), as defined in the DISTAL atlas, as a structure associated with ‘side-effects’. We presumed that VTAs covering at least an arbitrary 16 mm^3^ or 2000 voxels of the internal capsule would cause a side-effect and thus could not be used. As a result, this shifted the overall center of stimulation away from the neighboring internal capsule.

### Stimulation Scenarios

There are two typical scenarios occurring in previous studies that tried to determine DBS sweetspots. Many studies were based on the clinical outcome of clinically-used, chronic stimulation settings. These settings, derived over multiple clinical visits, should, at least in theory, lead to the individually best outcome in each patient. In such studies, there is typically only one stimulation setting (two hemispheres) which can be used for sweetspot mapping and thus larger patient cohorts are needed^9,12,13,15^. The second type of sweetspot mapping studies rely on stimulation data from acutely investigated stimulation settings, most often as part of so-called monopolar reviews. In these studies, multiple stimulation configurations per patient, often from each available electrode with increasing amplitudes are used. Such datasets naturally incorporate both; stimulation data with favorable, but also less desirable clinical outcome. Since a multitude of datapoints exist for each patient but stimulation testing can be burdensome, these studies often rely on smaller cohorts ^6,10,11,14^. For this study, we tried to reflect both scenarios in two different analyses.

For the first scenario (‘Clinical Scenario’) we selected only one of the calculated 20 VTAs per lead. In leads where no VTA caused an improvement of at least 50%, the VTA associated with the highest improvement was chosen. In leads in which multiple VTAs were associated with an improvement greater than 50%, the VTA with the highest ratio of improvement divided by stimulation amplitude was chosen., thus reflecting clinical programming where in general the stimulation setting with the lowest amplitude is chosen when multiple settings lead to a satisfactory improvement. Having selected one VTA for each of the 200 leads, we then randomly selected 100 leads/VTAs to reflect a typical cohort size for DBS mapping trials with long-term outcomes^7,9,12,13,15^.

For the second scenario (‘Monopolar Review Scenario’) we used all of the 20 VTAs per lead, reflecting data generated during a monopolar review. However we limited our analysis to 20 randomly selected leads, resulting in 400 VTAs, which again reflects a cohort size typical of mapping trials based on monopolar review data^6,11,14^.

Each of the two stimulation scenarios was investigated with only the sensorimotor STN or the sensorimotor STN + 1C, each with and without Gaussian noise applied to the clinical outcomes resulting in eight scenarios in total (Figure 1).

**Figure 1.**
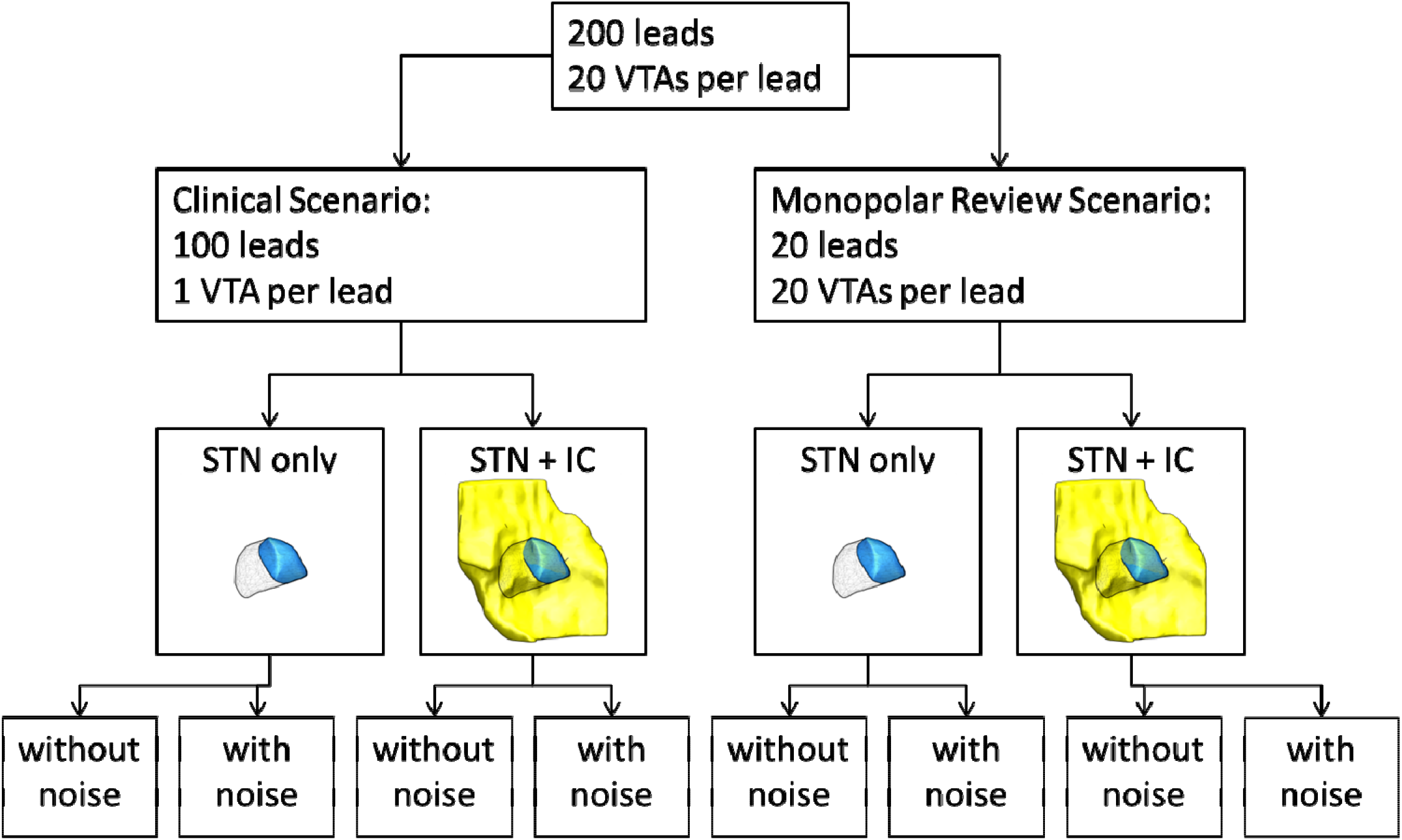
Overview of the investigated scenarios

### Sweetspot mapping

For each of these eight scenarios we investigated n = 5 sweetspot mapping methods which have been developed and evolved over the course of almost a decade. An overview of these methods can be found in Table 1 and a detailed description of each method is included as Supplementary Material 1. In chronological order, 1) Butson et al. first mapped a sweetspot in PD patients using a VTA-based mean-effect image which was thresholded to select voxels with an average clinical improvement > 50% to define a sweetspot.^10^ 2) Cheung et al. defined a sweetspot for dystonia by investigating which voxels were most often covered by VTAs in patients with good DBS outcome.^9^ From their cohort, they only used the VTAs of the 75 % of patients with the best outcome and then selected all voxels which were stimulated by at least 75% of these VTAs to define their sweetspot. In 2014, 3) Eisenstein et al. introduced voxel-wise statistics to the field of DBS mapping to define voxels associated with a significant improvement for different motor- and non-motor PD symptoms.^13^ Interestingly, their method was not based on VTAs but outcome was weighted for each voxel dependent on its distance to the active electrode. Then the resulting weighted outcomes for each voxel were tested against zero to determine voxels which lead to a significant improvement. They also addressed the problem of multiple comparisons inherent to voxel-wise statistics by expanding their method with a post-hoc nonparametric permutation statistic. 4) Reich et al. used a VTA based mean-effect image and again used voxel-wise statistics to identify voxels which had a significantly higher outcome than the average outcome of all VTAs which did not stimulate that particular voxel.^15^ Clusters of significant voxels were identified using a fixed threshold of n = 500 voxels for cluster size with smaller clusters being discarded. The same year, 5) our group (Dembek et al.) used a method in monopolar review data of PD patients.^6^ A mean-effect image was generated and voxel-wise statistics were used to compare the outcomes in each voxel against average, amplitude-corrected outcomes of the cohort as a whole. Based on Eisenstein et al. we then used a nonparametric permutation statistic to correct for multiple comparisons and identify valid clusters of significant voxels. For this study, these five mentioned methods were recreated in Matlab® (The Mathworks, Natick, USA). Other methods presented in the past were excluded from our analysis for being highly similar to either the method by Eisenstein et al.^12^, Dembek et al.^11^ or for using clinical data not comparable with our stimulation scenarios^14^. For all analyses, only voxels which were at least covered by n = 16 VTAs were regarded, to allow reliable statistics^6,11^.

**Table 1.**
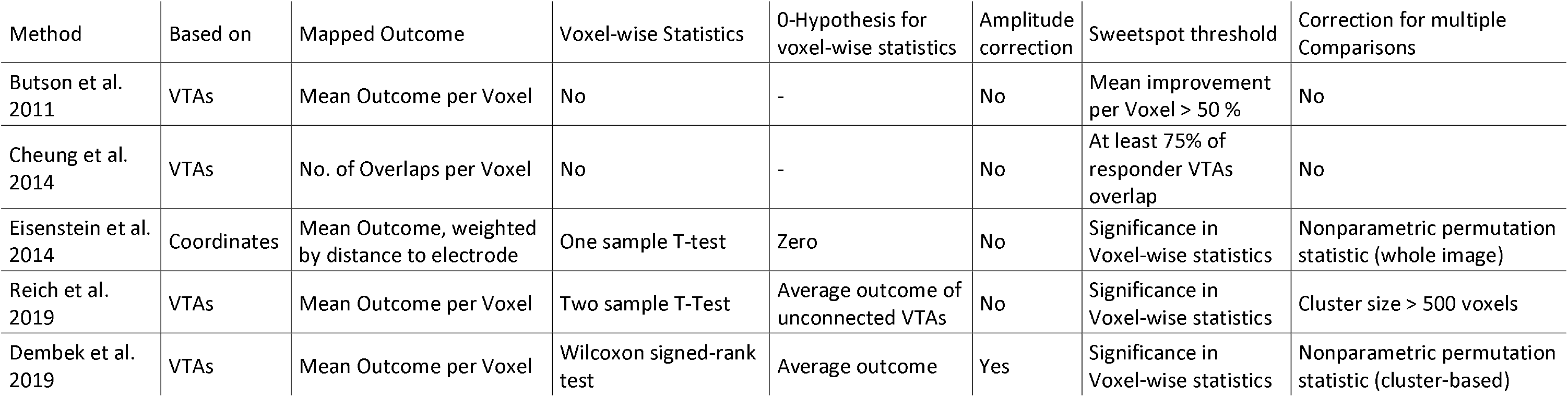
Overview of sweetspot mapping methods

| Method | Based on | Mapped Outcome | Voxel-wise Statistics | 0-Hypothesis for voxel-wise statistics | Amplitude correction | Sweetspot threshold | Correction for multiple Comparisons |
| --- | --- | --- | --- | --- | --- | --- | --- |
| Butson et al. 2011 | VTAs | Mean Outcome per Voxel | No | - | No | Mean improvement per Voxel > 50 % | No |
| Cheung et al. 2014 | VTAs | No. of Overlaps per Voxel | No | - | No | At least 75% of responder VTAs overlap | No |
| Eisenstein et al. 2014 | Coordinates | Mean Outcome, weighted by distance to electrode | One sample T-test | Zero | No | Significance in Voxel-wise statistics | Nonparametric permutation statistic (whole image) |
| Reich et al. 2019 | VTAs | Mean Outcome per Voxel | Two sample T-Test | Average outcome of unconnected VTAs | No | Significance in Voxel-wise statistics | Cluster size > 500 voxels |
| Dembek et al. 2019 | VTAs | Mean Outcome per Voxel | Wilcoxon signed-rank test | Average outcome | Yes | Significance in Voxel-wise statistics | Nonparametric permutation statistic (cluster-based) |

### Statistical analysis and outcomes

Each of the eight scenarios was repeated 30 times, each time using randomly selected DBS leads. For each of the 30 iterations we analyzed the following outcome parameters: To address the overall spatial concordance between the sensorimotor STN and the sweetspots detected by each method, we calculated the Sørensen-Dice-coefficient. The Sørensen-Dice-coefficient (Dice-coefficient for short) is determined by calculating two times the size of the overlap between two datasets and dividing this by the sum of the individual sizes of the two datasets. It thus ranges from Dice = 0, meaning no overlap between datasets, to Dice = 1, indicating perfectly congruent datasets. To assess the clinical relevance of the sweetspots, we also determined their predictive capabilities in out of sample data. Prediction was assessed by calculating the overlap between the sweetspot determined during one iteration with the VTAs from another randomly selected iteration. Then linear regression analysis was performed to determine what amount of variance (R^2^) in clinical outcome could be explained by the overlap with the sweetspot. Dice-coefficients and R^2^-values were compared between the five methods using a Friedman test with post-hoc pairwise comparisons using the Wilcoxon signed-rank test, Bonferroni-corrected for the number of comparisons (n = 10). Additionally, the sizes of sweetspots were calculated and compared to the size of the sensorimotor STN for each mapping method using Wilcoxon signed-rank tests, again Bonferroni-corrected for the number of comparisons (n = 5). All results are shown as medians ± range with Bonferroni-corrected p-values which were deemed significant if < 0.05.

### Further analyses

To describe the internal consistency of sweetspots generated by each method, we calculated Dice-coefficients between all sweetspots generated by one method during the 30 iterations for each scenario. Additionally, we calculated ‘average sweetspots’ for each method and each scenario, by determining which voxels were part of the respective sweetspot in at least 50 % of the 30 iterations.

## Results

### Clinical Scenario

Results for the Clinical Scenario are summarized in Figure 2 (STN) and Figure 3 (STN + 1C). The Friedman test revealed significant differences between methods for all analyses. Importantly, the post-hoc nonparametric permutation analysis of the Eisenstein et al. method discarded the detected sweetspots in scenarios where noise had been added. When comparing Dice-coefficients in our clinical scenario without added noise and without the internal capsule as an exclusion zone the Dembek et al. method significantly outperformed all other methods. When adding Gaussian noise to the clinical outcomes, the method by Cheung et al. had the highest Dice-coefficients. After excluding VTAs overlapping the internal capsule, the methods by Reich et al. and Dembek et al. both had significantly higher Dice-coefficients than other methods. The predictive capabilities, as measured by the amount of explained variance R^2^ were highest for the methods by Reich et al. and Dembek et al., which were able to explain around 80% of the variance in clinical outcome. The sweetspots determined by the Butson et al. and Eisenstein et al. methods were significantly larger than the sensorimotor STN, while the method by Cheung et al. consistently led to smaller sweetspots.

**Figure 2.**
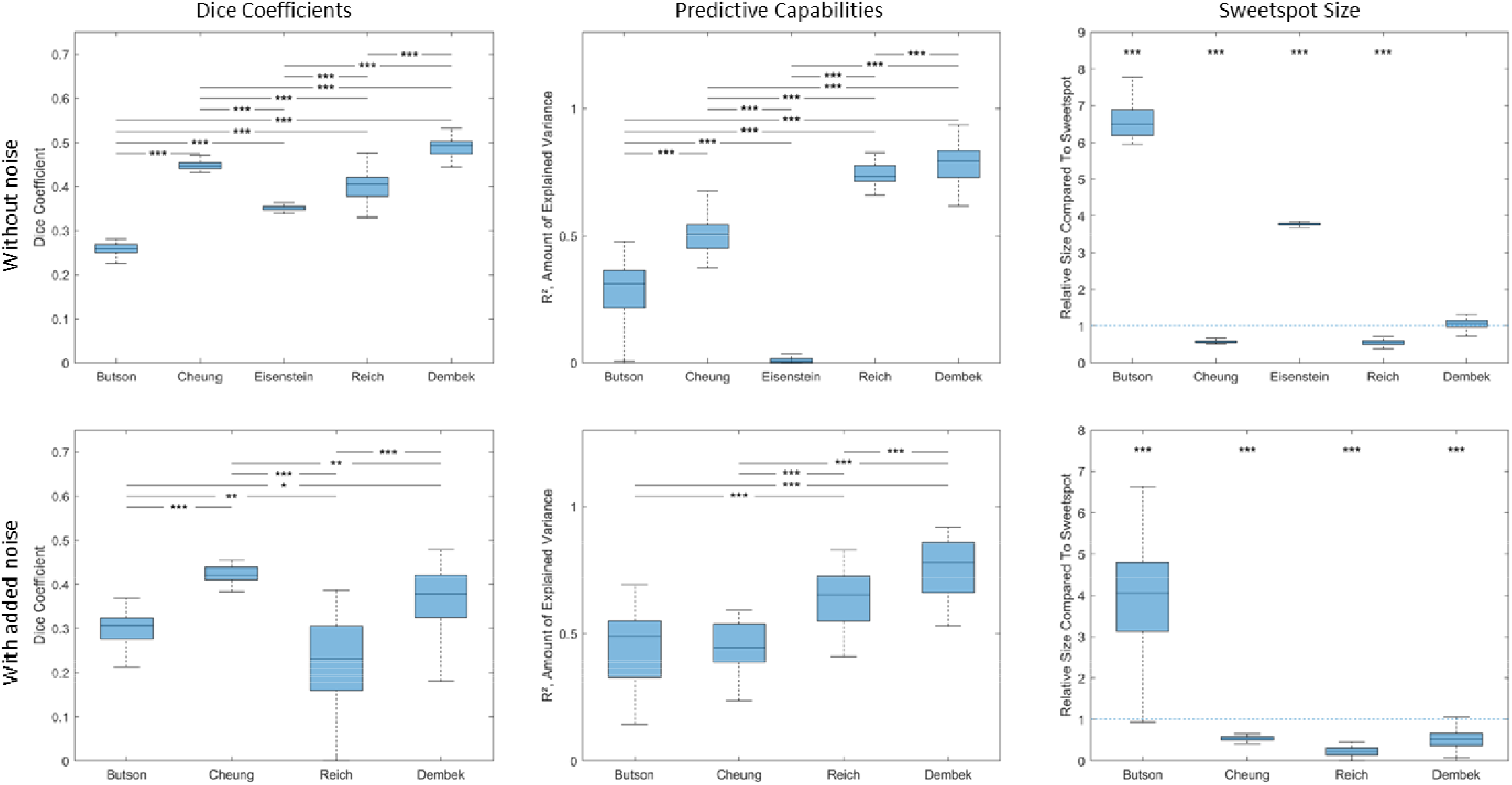
Clinical Scenario; STN

Legend: Results of the clinical scenario without the internal capsule, both without (first row) and with noise added to the clinical outcomes nd row). Boxplots are shown for Dice-coefficients (first column), the predictive capabilities (second column) and the relative sizes of tspots with respect to the sensorimotor STN (third column). Boxplots depict the median, interquartile range and range. The statistical kance of pairwise comparisons is indicated using asterisks for Dice-coefficients and the amount of explained variance. For size, the statistical kance between each method and the size of the sensorimotor STN are indicated using asterisks. Asterisks indicate significance for p-values cted for multiple comparisons: * p < 0.05; ** p < 0.01; *** p < 0.001.

**Figure 3.**
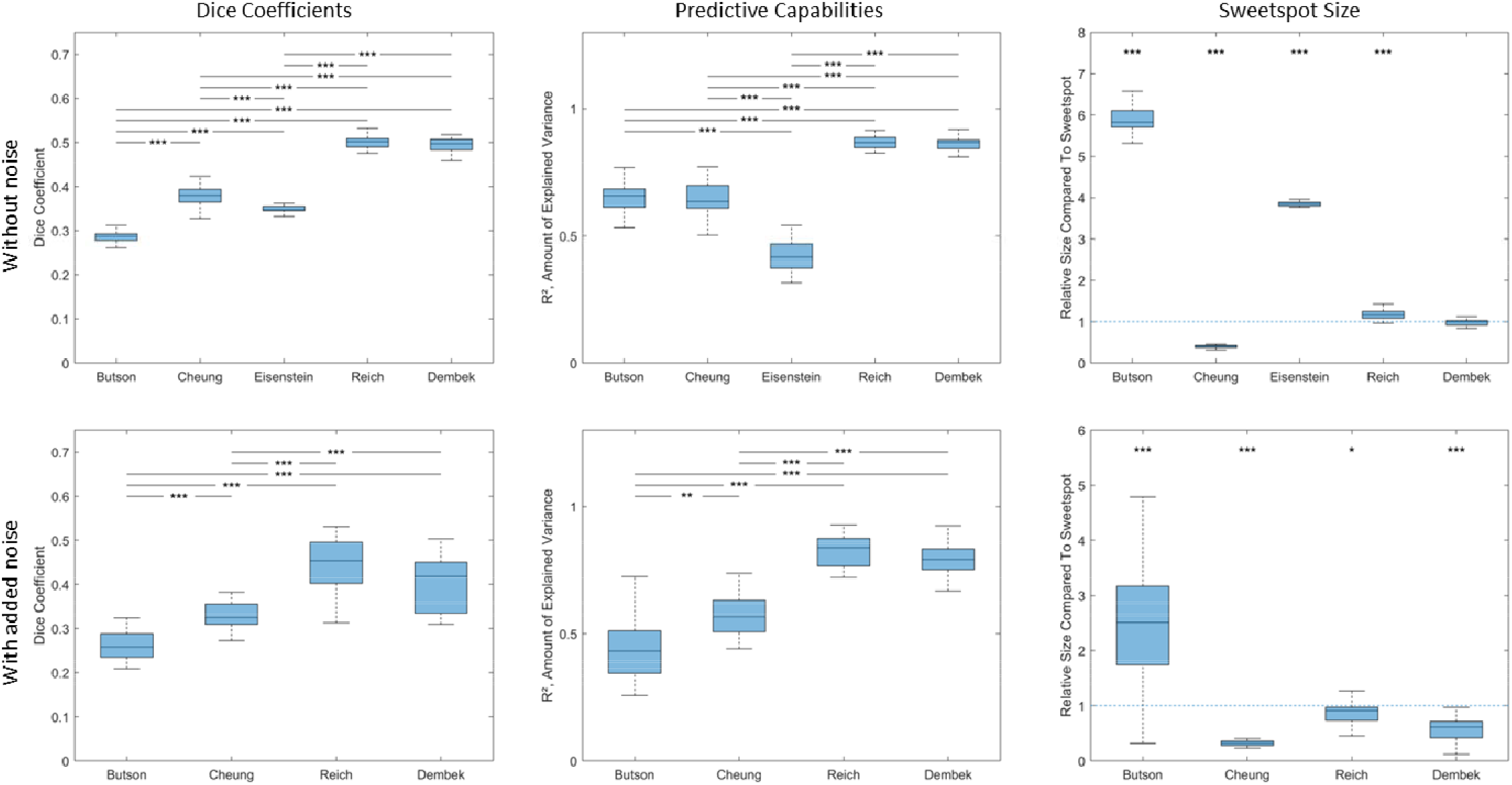
Clinical Scenario; STN + IC

Legend: Results of the clinical scenario with the internal capsule (1C), both without (first row) and with noise added to the clinical outcomes nd row). Boxplots are shown for Dice-coefficients (first column), the predictive capabilities (second column) and the relative sizes of tspots with respect to the sensorimotor STN (third column). Boxplots depict the median, interquartile range and range. The statistical kance of pairwise comparisons is indicated using asterisks for Dice-coefficients and the amount of explained variance. For size, the statistical kance between each method and the size of the sensorimotor STN are indicated using asterisks. Asterisks indicate significance for p-values cted for multiple comparisons: * p < 0.05; ** p < 0.01; *** p < 0.001.

### Monopolar Review Scenario

Results for the Monopolar Review Scenario are summarized in Figure 4 (STN) and Figure 5 (STN + 1C). Surprisingly, the method presented by Cheung et al. was not able to identify sweetspots in our data since the maximum number of overlaps of high outcome VTAs never reached the predefined 75% threshold. Also, sweetspots detected by the Eisenstein et al. method were again discarded during its nonparametric permutation analysis in scenarios with added noise. Dice-coefficients and predictive capabilities as described by the amount of explained variance R^2^ were significantly higher in the Dembek et al. method compared to all other remaining methods for all variants of the monopolar review scenario (with/without internal capsule; with/without noise). About 90% of variance in outcomes could be explained using these sweetspots. The Reich et al. method showed the second highest Dice-coefficients and predictive capabilities in all cases.

**Figure 4.**
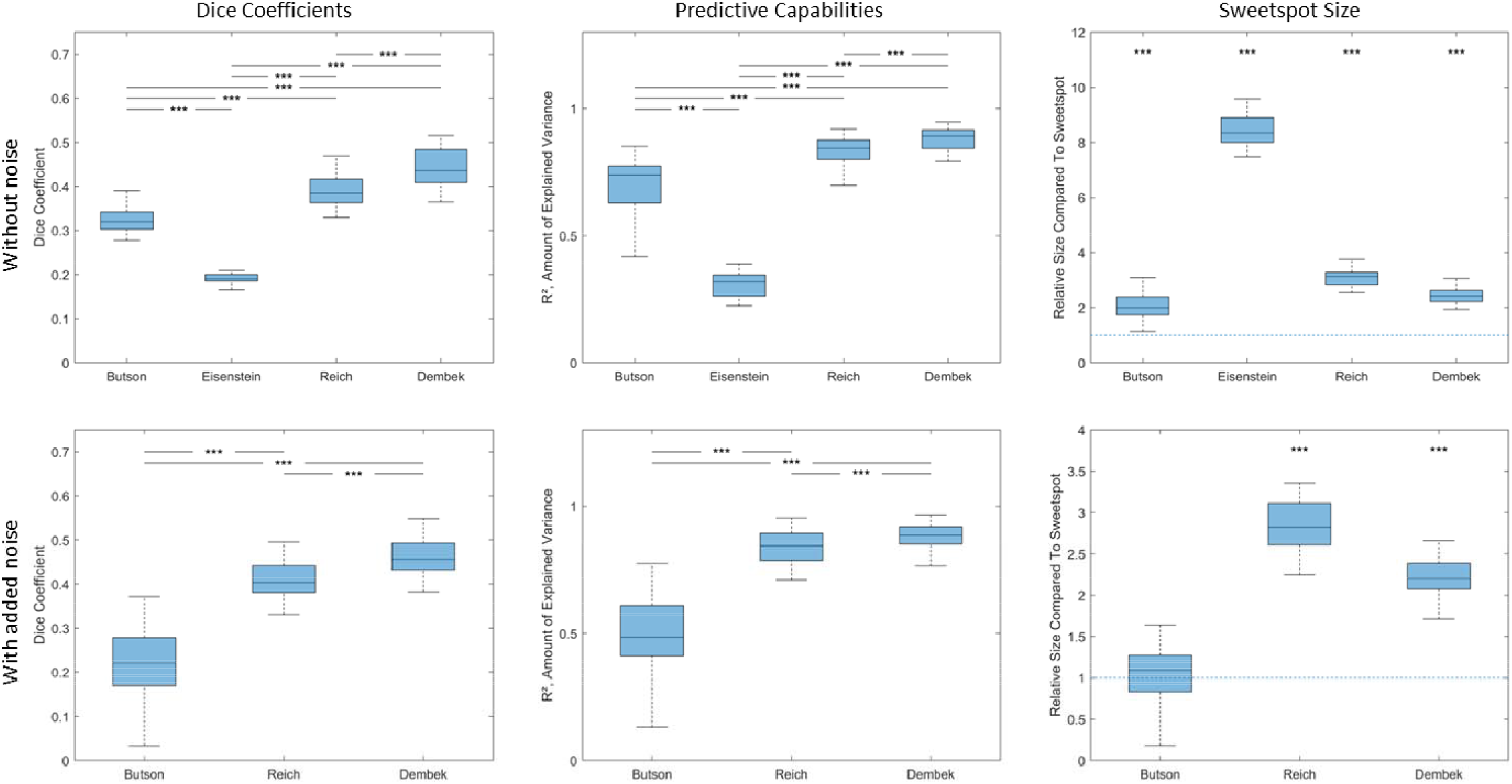
Monopolar Review Scenario; STN

Legend: Results of the monopolar review scenario without the internal capsule, both without (first row) and with noise added to the clinical imes (second row). Boxplots are shown for Dice-coefficients (first column), the predictive capabilities (second column) and the relative sizes *zee*tspots with respect to the sensorimotor STN (third column). Boxplots depict the median, interquartile range and range. The statistical kance of pairwise comparisons is indicated using asterisks for Dice-coefficients and the amount of explained variance. For size, the statistical kance between each method and the size of the sensorimotor STN are indicated using asterisks. Asterisks indicate significance for p-values cted for multiple comparisons: * p < 0.05; ** p < 0.01; *** p < 0.001.

**Figure 5.**
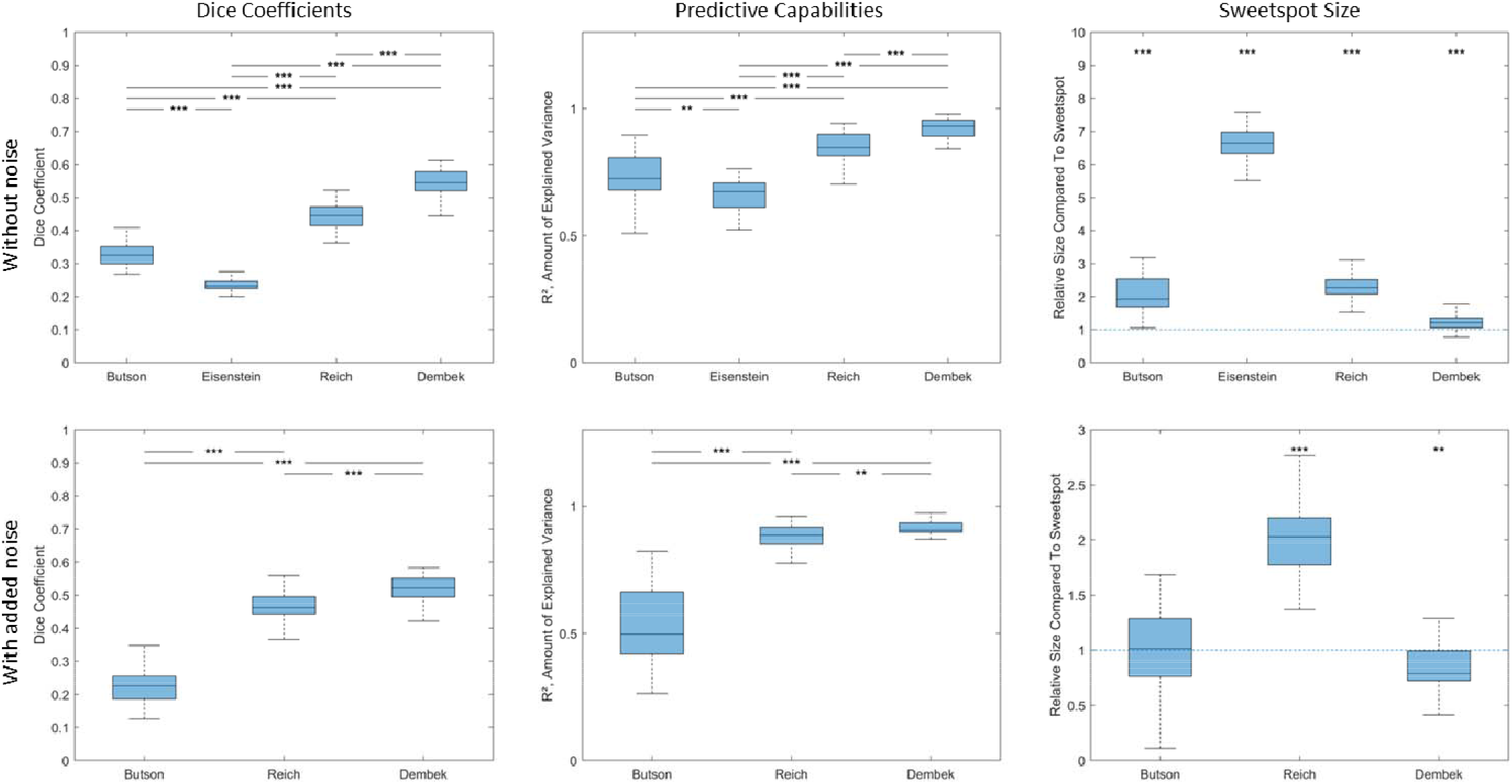
Monopolar Review Scenario; STN + IC

Legend: Results of the monopolar review scenario with the internal capsule (1C), both without (first row) and with noise added to the clinical imes (second row). Boxplots are shown for Dice-coefficients (first column), the predictive capabilities (second column) and the relative sizes *zee*tspots with respect to the sensorimotor STN (third column). Boxplots depict the median, interquartile range and range. The statistical kance of pairwise comparisons is indicated using asterisks for Dice-coefficients and the amount of explained variance. For size, the statistical kance between each method and the size of the sensorimotor STN are indicated using asterisks. Asterisks indicate significance for p-values cted for multiple comparisons: * p < 0.05; ** p < 0.01; *** p < 0.001.

### General sweetspot characteristics

Overall methods resulted in characteristic sweetspots which were often consistent within each method (see Figure 6 for average sweetspots and Table 2 for within-methods Dice-coefficients). The Butson et al. method, based on a 50% improvement threshold applied to the Mean-image often generated ‘shell-like’ sweetspots which emphasized on the posterior border of the investigated area. Results were quite inconsistent as illustrated by the reduced Dice-coefficient within the method, especially when noise was added. The Cheung et al. method, based on a fixed threshold applied to the N-image of responder VTAs resulted in very consistent, sphere-like sweetspots, which were close to the center of the overall investigated area and mostly lay within the sensorimotor STN. When using the Eisenstein et al. method with its voxel-wise statistics against zero, sweetspots tended to be large, round volumes, covering large parts of the investigated area. While there was a high consistency within the method for different iterations, the sweetspots lacked confinement to the actual sensorimotor STN. Reich et al. and Dembek et al. methods, which use voxel-wise statistics against average outcomes, resulted in quite similar types of sweetspots. Those were mainly centered on the sensorimotor STN with some overspill outside its medial and lateral border. Consistency within these methods sometimes proved to be an issue when noise was added.

**Table 2.** Dice-coefficients within methods

|  |  |  | Butson et al. | Cheung et al. | Eisenstein et al. | Reich et al. | Dembek et al. |
| --- | --- | --- | --- | --- | --- | --- | --- |
| Clinical<br>Scenario | STN | no noise | 0.89 (0.80-0.95) | 0.92 (0.79-0.96) | 0.96 (0.91-0.97) | 0.82 (0.50-0.91) | 0.83 (0.60-0.92) |
|  |  | with noise | 0.63 (0.07-0.88) | 0.91 (0.77-0.96) | - | 0.34 (0.00-0.83) | 0.48 (0.04-0.79) |
|  | STN + IC | no noise | 0.90 (0.80-0.96) | 0.88 (0.77-0.95) | 0.95 (0.92-0.98) | 0.88 (0.73-0.94) | 0.88 (0.77-0.93) |
|  |  | with noise | 0.33 (0.02-0.86) | 0.84 (0.66-0.94) | - | 0.74 (0.51-0.88) | 0.65 (0.39-0.86) |
| Monopolar<br>Review<br>Scenario | STN | no noise | 0.57 (0.29-0.90) | - | 0.87 (0.75-0.92) | 0.82 (0.65-0.92) | 0.81 (0.61-0.92) |
|  |  | with noise | 0.33 (0.06-0.77) | - | - | 0.81 (0.58-0.91) | 0.79 (0.55-0.91) |
|  | STN + IC | no noise | 0.57 (0.29-0.83) | - | 0.86 (0.70-0.93) | 0.79 (0.56-0.92) | 0.75 (0.42-0.90) |
|  |  | with noise | 0.27 (0.04-0.68) | - | - | 0.77 (0.52-0.89) | 0.67 (0.41-0.84) |

Legend: Median (range) Dice-coefficients within each of the five investigated methods (columns) and each of the eight scenarios (rows) are shown.

**Figure 6.**
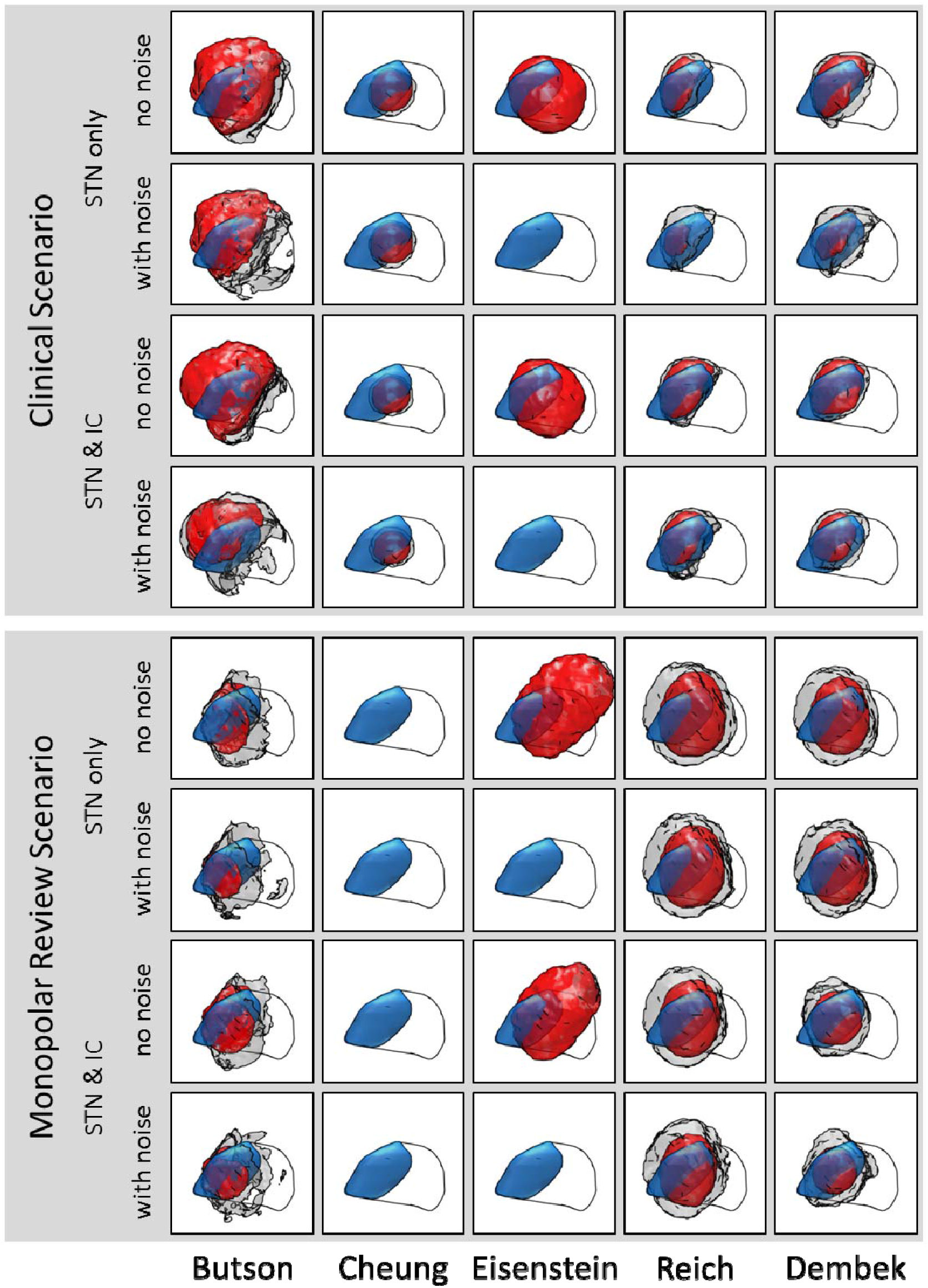
Average Sweetspots

Legend: Average sweetspots (red) signify all voxels localized within the sweetspot at least 50 *%* of times for the respective method (columns) and scenario (rows). Grey volumes on the other hand encompass all voxels which lay inside a sweetspot at least once. The sensorimotor STN (blue) signifies the ground-truth sweetspot. The whole STN is shown as black outline.

### Effects of post-hoc analyses

Three of the investigated methods, namely Eisenstein et al., Reich et al., and Dembek et al., used post-hoc analyses to correct for multiple voxel-wise testing (see Table 3 for detailed results). The Eisenstein et al. method, which evaluates the overall significance of the significant voxels against the overall significance of n = 200 permuted datasets discarded the whole sweetspot in all scenarios with added noise – and in none of the scenarios without added noise. The Reich et al. method, which discarded clusters of significant voxels whenever the size of the cluster was below n = 500 voxels mostly discarded scattered voxels and clusters consisting of fewer than 10 voxels. Such removal of scattered voxels mainly occurred in scenarios with added noise. The majority of those voxels was outside the ground-truth sweetspot. The Dembek et al. method, which ranked the overall significance of clusters of significant voxels against the most significant clusters of n = 1000 permuted datasets, also removed mostly small, scattered clusters. Cluster removal occurred more frequently than during the Reich et al. method, but again most removed voxels were outside the ground-truth sweetspot.

**Table 3.**
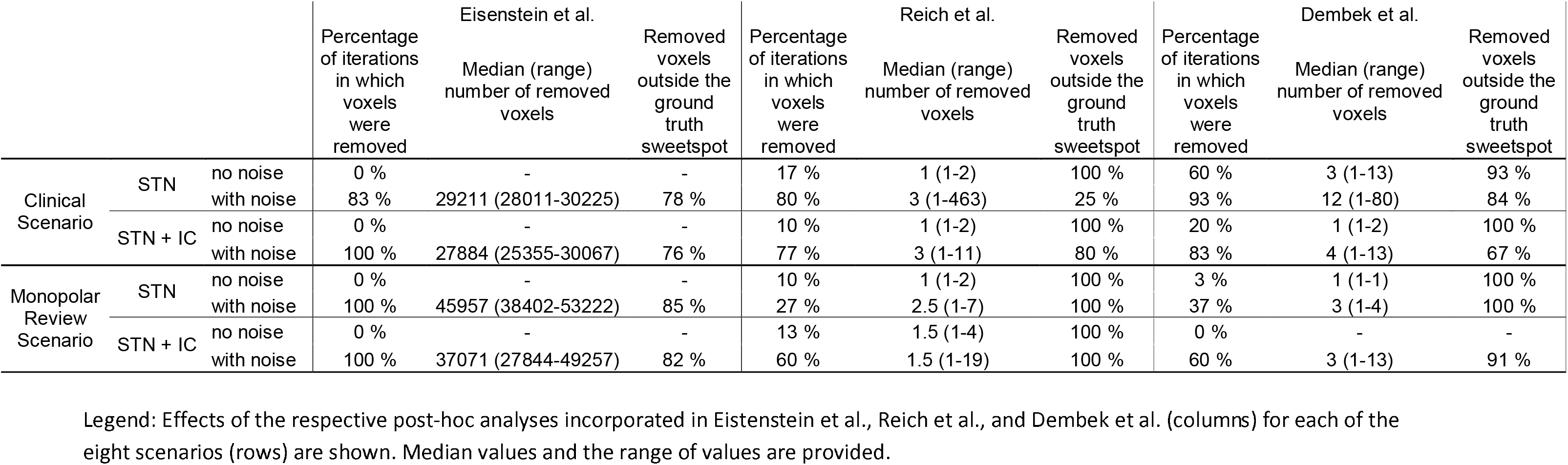
Effects of post-hoc analyses

|  |  |  | Eisenstein et al. |  |  | Reich et al. |  |  | Dembek et al. |  |  |
| --- | --- | --- | --- | --- | --- | --- | --- | --- | --- | --- | --- |
|  |  |  | Percentage of iterations in which voxels were removed | Median (range) number of removed voxels | Removed voxels outside the ground truth sweetspot | Percentage of iterations in which voxels were removed | Median (range) number of removed voxels | Removed voxels outside the ground truth sweetspot | Percentage of iterations in which voxels were removed | Median (range) number of removed voxels | Removed voxels outside the ground truth sweetspot |
| Clinical Scenario | STN | no noise | 0 % | - | - | 17 % | 1 (1-2) | 100 % | 60 % | 3 (1-13) | 93 % |
|  |  | with noise | 83 % | 29211 (28011-30225) | 78 % | 80 % | 3 (1-463) | 25 % | 93 % | 12 (1-80) | 84 % |
|  | STN + IC | no noise | 0 % | - | - | 10 % | 1 (1-2) | 100 % | 20 % | 1 (1-2) | 100 % |
|  |  | with noise | 100 % | 27884 (25355-30067) | 76 % | 77 % | 3 (1-11) | 80 % | 83 % | 4 (1-13) | 67 % |
| Monopolar Review Scenario | STN | no noise | 0 % | - | - | 10 % | 1 (1-2) | 100 % | 3 % | 1 (1-1) | 100 % |
|  |  | with noise | 100 % | 45957 (38402-53222) | 85 % | 27 % | 2.5 (1-7) | 100 % | 37 % | 3 (1-4) | 100 % |
|  | STN + IC | no noise | 0 % | - | - | 13 % | 1.5 (1-4) | 100 % | 0 % | - | - |
|  |  | with noise | 100 % | 37071 (27844-49257) | 82 % | 60 % | 1.5 (1-19) | 100 % | 60 % | 3 (1-13) | 91 % |

Legend: Effects of the respective post-hoc analyses incorporated in Eistenstein et al., Reich et al., and Dembek et al. (columns) for each of the eight scenarios (rows) are shown. Median values and the range of values are provided.

## Discussion

This study provides insight into the question of how different methods of DBS mapping influence the resulting sweetspots. Using ground-truth data, this study explores the possibilities and especially the limitations for different approaches. As the main finding, different previously published approaches results in a variety of different DBS sweetspots when used in the same dataset and results can diverge widely even within methods for different use-case scenarios.

The investigated methods were developed over almost a decade, with newer methods heavily relying on their forebears. As a result of this evolution, we indeed found that the most recent approaches by Reich et al.^15^ and Dembek et al.^6^, which employed voxel-wise statistics comparing the outcomes of each voxel against an average of other outcomes in the dataset, provided the best results. Overall, these methods provided sweetspots with the highest Dice-coefficients (i.e. similarity between computed and actual ground-truth sweetspot) and the highest predictive capabilities. A correction for stimulation amplitude, as implemented in Dembek et al.^6^, seemed to further improve results, especially with monopolar review data in which amplitude naturally plays a major role. Post-hoc analyses i. e. cluster thresholding or nonparametric permutation statistics helped to remove some scattered clusters of voxels but did not affect the results in this study in a major way. In real-life data however, false positives may play a larger role and post-hoc correction thus might be more important.

Employing a voxel-wise statistical analysis against zero, as implemented in the Eisenstein et al.^13^ method, consistently led to large parts of the investigated area being highlighted as significant. This is not surprising due to the nature of our data which only included clinical improvements and not clinical worsening. However, worsening of e.g. motor symptoms due to DBS is also rarely observed in clinical reality, so assuming zero improvement as the null-hypothesis for voxel-wise statistical analysis indeed might be too liberal and will provide less meaningful clinical insights. This reasoning is supported by the low amount of explained variance in our prediction analysis and the fact that the post-hoc analysis implemented in the method itself removed a lot of the resulting sweetspots as possible false-positives^11^.

The approach by Cheung et al.^9^, which investigated where most responder VTAs overlap, provided relatively consistent results in clinical data which remained comparably unaffected by the addition of noise. Of note, this method underestimated the size of the sweetspot. Moreover, since this approach tended severely towards the overall center of stimulation, it performed worse when the center of stimulation was moved away from the sweetspot, e.g. in the scenarios with the added internal capsule. Additionally, this approach did not work with monopolar review data – at least not when using the predefined overlap threshold of 75%.

Importantly, the maximum achievable Dice-coefficients even in the best cases were around 0.5, meaning that there was still a significant mismatch between the calculated sweetspots and the anatomical ground-truth. This result highlights the general limitations of current approaches to DBS sweetspot mapping regarding neuroanatomical accuracy. These limitations become even more important when one considers that the many other sources of noise present in clinical reality, like e.g. imaging-inaccuracies, uncertainties regarding the VTA-model, or the largely unknown underlying electric tissue properties, did not factor into this *in silico* analysis. Additionally, one has to keep in mind that as a general limitation only those parts of a sweetspot can be identified which are covered by enough stimulation data. In reality, it might of course happen that the ‘real sweetspot’ is so far off target, that it cannot be discovered at all using sweetspot mapping.

On the other hand, the Dembek et al. and Reich et al. methods could explain large amounts of variance during the out-of-sample prediction analysis – highlighting their potential use in future applications like computer-guided DBS programming. Taken together, the mismatch between suboptimal anatomical accuracy and high predictive capabilities also sheds light onto another important fact: namely that even if a sweetspot is able to predict clinical outcome, it does not mean that its anatomical location is necessarily linked to the underlying neuroanatomical structure causing the clinical effect. This is of particular importance when drawing conclusions about the underlying neuroanatomy or networks from such analyses^7,8,11,26–28^.

## Towards an improved DBS mapping strategy

When considering the results of this study as well as the many possible sources of noise in DBS sweetspot mapping, it seems quite difficult to develop a strategy which can sufficiently address all these factors. In our opinion, the ideal mapping strategy should not rely on fixed and arbitrary thresholds. As demonstrated in this study, methods based on arbitrary thresholds are highly sensitive to differences in use-case scenarios. Even if such thresholds were defined in ground-truth models, one could never be sure that these models genuinely reflect clinical reality. Voxel-wise statistics investigating outcome in a certain voxel against (average) clinical outcomes in other DBS settings thus seem to be the most promising approach at the moment. Such methods do not rely on thresholding and, as shown in our analyses, seem to provide relatively consistent results for different scenarios. A possible avenue for further improvement might be using more advanced statistical methods for voxel-wise testing. Instead of t-tests^12,13,15^ or Wilcoxon signed-rank tests^6,11^, voxel-wise mixed-effect models might for example be a promising strategy to account for clinical covariates like symptom severity or medication. They could also help to expand the relatively narrow scope of the ‘VTA’ itself by incorporating other factors like stimulation frequency or pulse-width, different stimulation configurations like anodal, bipolar or multi-electrode stimulation^29,30^, or axon properties like activation thresholds or directionality. Regarding post-hoc corrections for multiple comparisons, we again think that threshold-based methods like cluster thresholding should be avoided. While of course being highly inefficient from a computational point of view, nonparametric permutation statistics have the important advantage of being comparatively free of underlying assumptions. Another important question is whether binary sweetspots are the optimal solution for describing areas with good stimulation outcome. By binarizing DBS mapping results into a however well-defined sweetspot, a lot of the information about spatial gradients/contrasts is lost. While binarized sweetspots are of course useful and easily understandable approximations, it remains to be seen if more complex metrics might be better suited for e.g. outcome prediction analyses^15^.

No matter what methods will be developed and employed in the future, this study hopefully highlights the importance of ground-truth, *in silico* testing. Many previous DBS sweetspot studies have reached a broad audience, directly influencing our assumptions of where optimal stimulation outcome might be achieved. They thus have the potential to impact both: electrode placement and postoperative DBS programming. This highlights the importance of first examining a method’s viability in ground-truth data before using this method to draw clinically relevant conclusions. The same also holds true for new approaches to DBS mapping which incorporate for example functional and/or structural connectomic data^7,26,31^.

## Conclusion

The abilities of current DBS mapping methods to reliably detect sweetspots for motor outcomes are limited and potentially influenced by a variety of factors. While methods have evolved over the years, there is still much room for improvement. In silico validation should be used before drawing clinical conclusions from new DBS mapping methods and researchers should be aware of the respective strengths and limitations of each method.

## Data Availability

All data and scripts of this in silico trial can be made available upon request.

## Notes

Conflict of Interests: There were no conflicts of interest regarding this manuscript. Full disclosures are included for each author in the respective form.

### Competing Interest Statement

The authors have declared no competing interest.

### Funding Statement

There was no external Funding.

### Author Declarations

No ethics vote required.

